# Forecasting COVID-19 cases and deaths in epidemic-mitigating European countries by Richards function-based regression analyses

**DOI:** 10.1101/2020.05.18.20106146

**Authors:** Cheng Long, Xinmiao Fu

## Abstract

The COVID-19 pandemic has hit many countries, and in some European countries it has been mitigated since April. Here we applied Richards function to simulate and forecast the course of COVID-19 epidemics in Italy, Spain, France, Germany, Turkey, Belgium, Ireland, Netherlands, Portugal and Switzerland. Potential total COVID-19 confirmed cases in these countries were estimated to be 240400±1300, 294100±4000, 178500±800, 176900±700, 155400±1000, 57900±400, 24000±200, 46200±300, 30000±300 and 30700±100 respectively. Most of these countries are predicted to approach ending stage between late May and early June such that daily new cases will become minimal, which may guide societal and economic restorations. In addition, total COVID-19 deaths were estimated to be 33500 ±300, 28200±200, 27800±200, 8740±80, 4500 ±30, 9250±70, 1530±20, 6240±50, 1380±10 and 1960±8, respectively. To our best knowledge, this is the first study forecasting the COVID-19 epidemic by applying the Richard function-based regression analysis.

## Introduction

The COVID-19 pandemic caused by the novel coronavirus (SARS-CoV-2) has hit many countries [1], resulting in more than 4 million cases and over 300,000 deaths in the world as of May 14, 2020. In European countries such as Italy and Spain, daily new cases are declining since April, yet they are still as many as one thousand per day. In these epidemic-mitigating countries, governments are attempting to restore normal societal and economic activity, however, concerns are raised about the risk of COVID-19 resurgence. It is of significance to precisely forecast when the epidemic in each country will become minimal such that societal and economic restorations will not lead in the resurgence of the outbreak. Further, such forecasting may also inform governments to get better preparedness for making control measures and supplying medical resources. Estimations by traditional epidemiological models apparently require much detailed data for analysis [2, 3]. Here we applied a simple data-driven, Richards function-based approach for epidemic forecasting only based on the daily cumulative confirmed COVID-19 cases. This approach has been explored for simulating 2003 SARS outbreaks in several regions/countries by others [4, 5] and also for estimating COVID-19 deaths in China by us recently [6].

## Methods

The Richards function is expressed as follows [4, 5]:

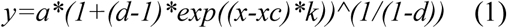

where *y* is the daily cumulative number of confirmed COVID-19 cases and *x* is day; *a*, *d, xc* and *k* are constants. Specifically, total number of COVID-19 cases are given by parameter *a* in the equation. Simulation was performed using Microcal Origin software with the Richard function.

## Results

### Estimation of potential total COVID-19 confirmed cases

We decided to collect the data of the cumulative confirmed cases from the website of Worldometer [7] for 10 most affected and epidemic-mitigating European countries, i.e., Italy, Spain, France, Germany, Turkey, Belgium, Ireland, Netherlands, Portugal and Switzerland (for detail, refer to the supporting information file **original data.xlsx**). Regression analyses indicate that the data of each country were well fitted with the Richards function (all *R^2^* values being close to 0.999; **Table 1** and **Figure 1**). Potential total numbers of confirmed cases in the above countries were estimated to be 240400±1300, 294100±4000, 178500±800, 176900±700, 155400±1000, 57900±400, 24000±200, 46200±300, 30000±300 and 30700±100 respectively. All parameters from regression analyses are presented in **Table S1** and can be utilized to predict daily new cases in each country since May 15 according to equation 1.

**Table 1.** Estimation of potential total COVID-19 confirmed cases and deaths in European countries.

| Countries | Total confirmed cases |  | Key date <sup>a</sup> | Total deaths |  | Crude fatality ratio (%) <sup>b</sup> |
| --- | --- | --- | --- | --- | --- | --- |
| | Number | $R^2$ | | Number | $R^2$ | |
| Italy | 240400±1300 | 0.999 | June 6 | 33500 ±300 | 0.999 | 13.9±0.9 |
| Spain | 294100±4000 | 0.999 | June 13 | 28200±200 | 0.999 | 9.6±0.9 |
| France | 178500±800 | 0.999 | May 18 | 27800±200 | 0.999 | 15.6±0.7 |
| Germany | 176900±700 | 0.999 | May 24 | 8740±80 | 0.999 | 4.9±0.9 |
| Turkey | 155400±1000 | 0.999 | June 8 | 4500 ±30 | 0.999 | 2.9±0.7 |
| Belgium | 57900±400 | 0.999 | June 5 | 9250±70 | 0.999 | 16.0±0.9 |
| Ireland, | 24000±200 | 0.998 | May 25 | 1530±20 | 0.997 | 6.4±1.3 |
| Netherlands | 46200±300 | 0.999 | June 4 | 6240±50 | 0.999 | 13.5±0.9 |
| Portugal | 30000±300 | 0.999 | June 9 | 1380±10 | 0.999 | 4.6±0.8 |
| Switzerland | 30700±100 | 0.999 | May 14 | 1960±8 | 0.999 | 6.4±0.4 |

<sup>a</sup> Key date is determined when daily new cases are less than 0.1% of the potential total confirmed cases.
<sup>b</sup> Crude fatality ratio was calculated as total deaths among total confirmed cases, and the standard deviation was calculated according to partial differential equations.

**Figure 1.**
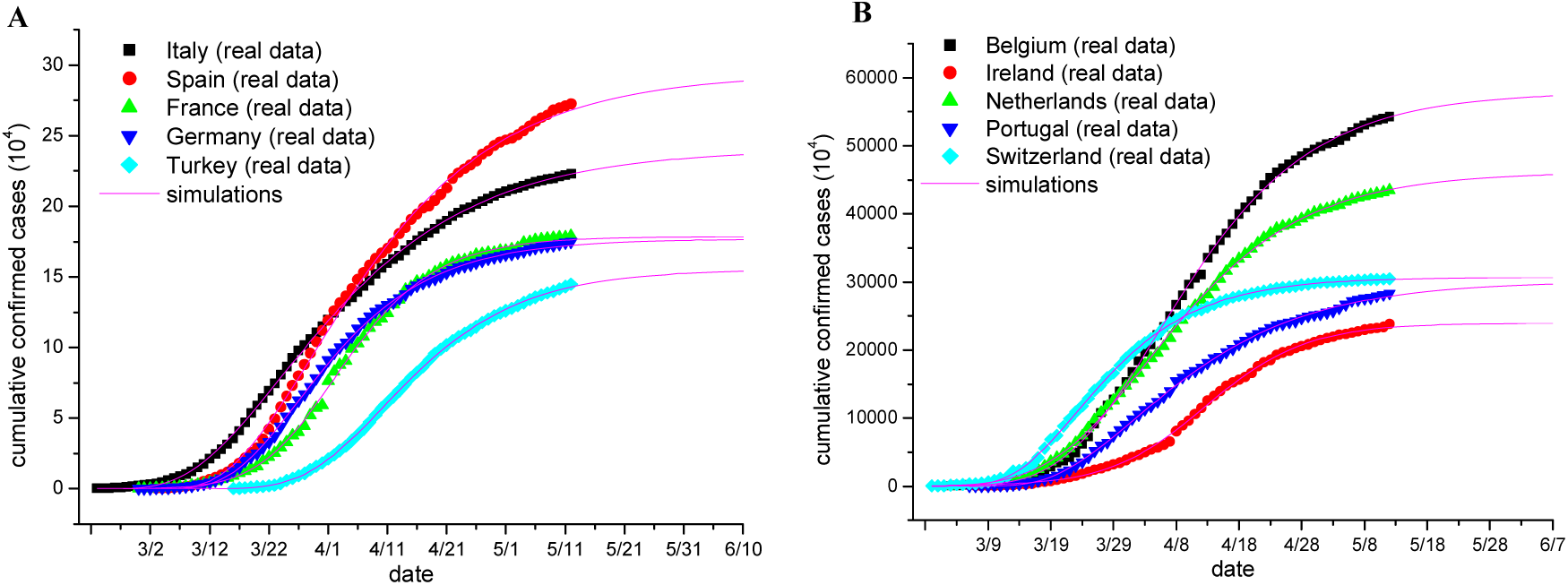
Fitting the cumulative confirmed COVID-19 cases of each European country to the Richards function. (**A, B**) Plots of the cumulative number of COVID-19 confirmed cases of the indicated European countries as of May 14, 2020, with the simulation results being plotted as Magenta lines. Simulation was performed using Microcal Origin software with the Richard function. All parameters from regression analyses are shown in Table S1.

In addition, we estimated the key date, on which daily new cases are lower than 0.1% of the potential total confirmed cases as defined by us subjectively. It appears that most countries will approach such late stage between late May and early June (refer to **Table 1**) such that daily new cases will become minimal in comparison with potential total confirmed cases. In our opinion, nationwide societal and economic restorations should be initiated at least not earlier than the estimated key dates, although local restorations may be operated earlier according to the local epidemic situation.

### Estimation of potential total COVID-19 deaths

In several countries, crude fatality ratios are as high as over 10% such that people have raised grave concerns about how many patients will die eventually. Assuming that the number of deaths is proportional to the number of confirmed cases for the COVID-19 outbreak under specific circumstances, we speculated that the cumulative COVID-19 deaths would also obey the Richards function. Regression analyses indicate that the data of cumulative COVID-19 deaths in each country were well fitted with the Richards function (all *R* values being close to 0.999; **Table 1** and **Figure 2**). All parameters of regression analyses are displayed in **Table S2** and can be utilized to predict daily new deaths in each country since May 15 according to equation 1. Potential total COVID-19 deaths in Italy, Spain, France, Germany, Turkey, Belgium, Ireland, Netherlands, Portugal and Switzerland were estimated to be 33500 ±300, 28200±200, 27800±200, 8740±80, 4500 ±30, 9250±70, 1530±20, 6240±50, 1380±10 and 1960±8, respectively. Crude fatality ratios (total deaths among total confirmed cases) of these countries could be estimated to be 13.9±0.9%, 9.6±0.9%, 15.6±0.7%, 4.9±0.9%. 2.9±0.7%, 16.0±0.9%, 6.4±1.3%, 13.5±0.9%, 4.6±0.8% and 6.4±0.4%, respectively.

**Figure 2.**
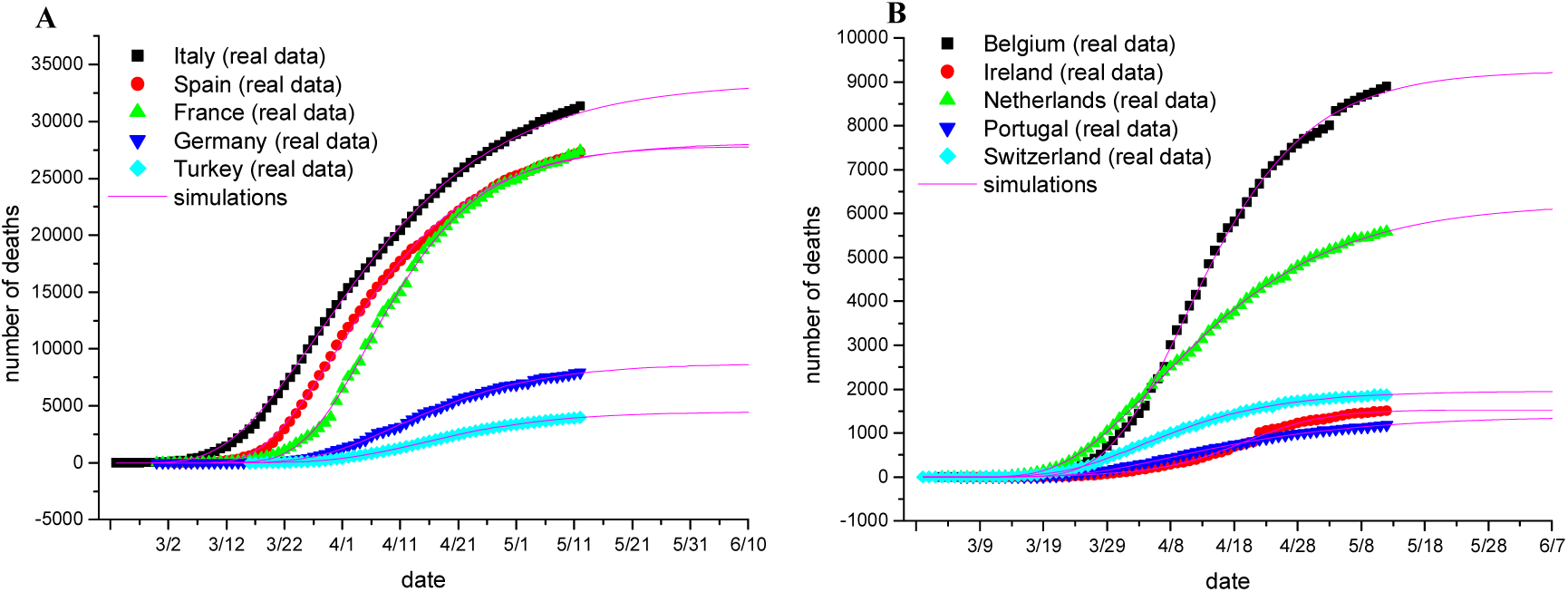
Fitting the cumulative COVID-19 deaths of each European country to the Richards function. (**A, B**) Plots of the cumulative number of COVID-19 confirmed cases of the indicated European countries as of May 14, 2020, with the simulation results being plotted as Magenta lines. Simulation was performed using Microcal Origin software with the Richard function. All parameters from regression analyses are shown in Table S2.

## Discussion

To our best knowledge, this is the first study forecasting the COVID-19 epidemic by applying the Richard function-based regression analysis. It should be pointed out that all the above estimates are based on the assumption that there are no significant changes regarding the current interventions, including non-pharmacological interventions such as quarantine and other prevention measures and also diagnosis and treatment approaches. In addition, UK and Sweden, two most affected European countries with more than 240,000 and 29,000 COVID-19 confirmed cases as of May 15, respectively, were not subjected to analysis because their daily new cases have not yet started to significantly decline (**Figure. S1**).

Collectively, our forecasting on the course of COVID-19 outbreaks in these European countries may provide valuable guidance for governments and also the public to get better preparedness and optimize their efforts to contain this unprecedented crisis at current critical stage. The estimated key date for each country may serve as a reference for operating societal and economic restorations such that the resurgence of COVID-19 can be avoided, minimized or under control according to COVID-19 containment in China [8, 9]. In addition, because thousands of COVID-19 patients have died in these countries, our prediction on the course of COVID-19 deaths (refer to **Figure 2** and **Table S2**) may benefit the mental health service that needs to be timely provided to the families of passed patients [10]. As a matter of fact, we precisely estimated total COVID-19 deaths in different areas of China (including Hubei Province and Wuhan city, the epicenter of the COVID-19 outbreaks in China [1]) by applying both Boltzmann and Richards functions-based regression analyses [6]. Such estimation help governments to evaluate the severity of the COVID-19 epidemics.

## Data Availability

Data are included in the manuscript

## Acknowledgments

This work is support by the National Natural Science Foundation of China (No. 31972918 and 31770830 to XF). We thank graduate students (Zuqin Zhang, Youhui Zeng, Shuting Shi and Zhijie Huang, all from Professor Fu’s research group at Fujian Normal University) for data collection.

## Author contribution

X.F. designed the study; C.L. organized data and performed analysis; X.F. wrote the manuscript.

## Conflict of interest

none.

## Notes

### Competing Interest Statement

The authors have declared no competing interest.

